# Adherence to the Eatwell Guide and associations with markers of adiposity: A prospective analysis within the UK Biobank cohort

**DOI:** 10.1101/2025.11.27.25341158

**Authors:** Alex Griffiths, Sarah Gregory, Fiona C Malcomson, Marie Spreckley, Jamie Matu, Louisa Ells, Oliver M Shannon

## Abstract

**Background:** Obesity remains a major public health concern in the UK, contributing towards increased disease risk and premature mortality. The Eatwell Guide - the UK’s health eating model - is widely applied in policy and practice, yet evidence linking adherence to this dietary pattern with adiposity is limited. Similarly, understanding whether associations differ across population subgroups, including by level of genetic risk for obesity, is essential to inform equitable and effective dietary guidance.

**Methods:** In 156 764 participants from the UK Biobank, we explored cross-sectional and prospective associations between adherence to the Eatwell Guide and markers of adiposity (BMI, waist circumference, a body shape index [ABSI], and total and trunk body fat percentage). Differences between population sub-groups including by genetic risk, age, sex, physical activity level and socioeconomic status were explored.

**Results:** Higher Eatwell Guide adherence was cross-sectionally associated with lower BMI (β = −0.032, SE = 0.001, p<0.001), with higher adherence associated with 25% lower odds of overweight/obesity versus lower adherence (OR = 0.75, 95% CI 0.73-0.77, p<0.001). Prospectively, greater Eatwell Guide adherence predicted more favourable BMI trajectories over time (β = −0.008, SE = 0.001, p<0.001). Similar, significant associations were observed for waist circumference, ABSI, and total and trunk body fat percentage (all p<0.05) and were broadly consistent across key population sub-groups.

**Conclusions:** Higher Eatwell Guide adherence was associated with beneficial changes in multiple markers of adiposity over time. These associations were consistent across key demographic groups, highlighting the potential role of adhering to UK healthy eating recommendations as part of weight management strategies in the UK.

## Introduction

Obesity remains a public health priority in the UK, with 28% and 36% of adults in England classified as having overweight or obesity in 2022 [1]. As such, around two-thirds of the adult population are at an increased risk of adverse, obesity-related health conditions such as cardiovascular disease [2], certain cancers [3], and premature mortality [4]. The substantial health and economic burden associated with obesity highlights the need for investigation into optimal dietary patterns for weight management, although this must be considered within a broader, person-centred approach that considers social, cultural, emotional and environmental factors [5]. Whilst emerging pharmacological interventions such as glucagon-like peptide-1 receptor agonists represent important advances in obesity treatment [6,7], understanding the role of diet quality remains essential for sustainable weight management and overall health [8].

The UK healthy eating recommendations are visually represented and communicated in a public facing tool called the Eatwell Guide [9]. The Eatwell Guide displays a dietary pattern which is characterised by consumption of a variety of fruits and vegetables, moderate consumption of starchy carbohydrates and wholegrains, moderate consumption of beans and pulses, meat, fish and eggs, adequate fluid intake, and low intake of foods rich in fat, salt and sugar [9,10]. Crucially, the Eatwell Guide is designed based on foods familiar and available to the UK population, which could help with uptake and adherence. Nevertheless, despite its prominent role in UK nutrition policy and practice, evidence linking Eatwell Guide adherence with health outcomes remains limited.

Higher adherence has been associated with reduced all-cause mortality across multiple UK cohorts [11], as well as an increase in life expectancy of ∼10 years in the UK Biobank cohort [12]. Cross-sectional improvements in cardiometabolic outcomes such as systolic and diastolic blood pressure, as well as a lower BMI, have been observed with greater Eatwell Guide adherence [13]. Notably, one prospective study in post-menopausal women found that higher Eatwell Guide adherence was associated with smaller increases in waist circumference over 4 years and reduced risk of abdominal obesity, although no changes in weight were observed [14]. Despite this, the current evidence base exploring Eatwell Guide adherence and markers of adiposity has several limitations. Existing research has primarily been conducted in relatively small cohorts with specific demographic characteristics. Cross-sectional work has focused on small (n=517) and specialised populations [13], whilst the only prospective study examined these associations only in post-menopausal women (n=4,162) [14]. These sample sizes and narrow demographics limit the ability to generalise to the wider UK population and to conduct adequately powered subgroup analyses. However, such analyses could identify demographic subgroups who may benefit most from adhering to the Eatwell Guide, thereby informing more targeted and effective public health interventions. In addition, whilst BMI and waist circumference are useful population level tools [15], they cannot comprehensively quantify body composition. Additional metrics which quantify both total and regional adiposity are required to provide complementary data which may better predict disease risk [16]. Finally, genetic predisposition has a key role in the development of obesity [17], yet no study to date has explored whether the associations between Eatwell Guide adherence and markers of adiposity are moderated by genetic risk. Understanding these interactions could clarify whether individuals at high genetic risk derive similar benefits from adhering to healthy dietary patterns as those at lower genetic risk.

The present study therefore aims to evaluate both cross-sectional and prospective associations between adherence to the Eatwell Guide and multiple markers of adiposity within the UK Biobank cohort. Specifically, we will investigate relationships between Eatwell Guide adherence and BMI, waist circumference, a body shape index (ABSI), total body fat percentage and trunk fat percentage at baseline, as well as changes in these measures over time. These findings will contribute to the evidence base evaluating UK healthy eating guidelines and may help to provide data on the suitability of the Eatwell Guide in supporting weight management and reducing the prevalence of obesity.

## Methods

### Study population and design

Prospective data were obtained from the UK Biobank, a cohort study of over 500 000 adults aged 37-73 years recruited from England, Scotland, and Wales, designed to investigate the determinants of disease in the UK population [18]. Full details have been published previously [19], but at baseline, participants completed a touchscreen questionnaire covering sociodemographic factors, diet, physical activity, and general health, as well as verbal interviews, measures of physical function and biological samples were provided. The current study included participants of any age, sex or ethnicity, with appropriate dietary data, a pre-specified marker of adiposity, and complete covariate data (see Statistical Analysis). Participants classified as underweight (BMI <18.5 kg/m²) were excluded due to physiological, medical and behavioural complexities that can affect the relationship between dietary intake and body weight in this population [20,21]. Ethical approval for the UK Biobank study was provided by the North West Haydock Research Ethics Committee (REC reference: 16/NW/0274), and all participants provided electronic signed consent.

### Dietary assessment and calculation of Eatwell Guide scores

In the current project, we used dietary data from the Oxford WebQ, a web-based, self-administered 24-hour dietary recall tool validated for use in large-scale epidemiological studies [22,23]. The Oxford WebQ captures information on the consumption of 206 food items and 32 beverages from the previous day, with participants indicating the number of standard portions consumed for each item. Participants completed the Oxford WebQ during their baseline assessment centre visit. Those who provided a valid email address were subsequently invited to complete up to four additional Oxford WebQ assessments approximately every three to four months. All available dietary assessments for each participant were used to assess adherence to the Eatwell Guide. Where multiple recalls were available these were averaged. Dietary recalls that participants identified as atypical were excluded from the analyses.

### Eatwell Guide adherence score

Eatwell Guide adherence was assessed using a graded, food-based scoring system developed and described in detail previously [24]. The approach focuses on foods rather than nutrients to align with the Eatwell Guide framework [9,10]. This approach offers more actionable guidance for public health messaging and allows researchers and participants to better quantify and track changes in diet over time [25].

The scoring tool (Supplementary Table S1) included components for starchy carbohydrates, wholegrains, beans and pulses, fish, white meat, nuts, eggs, fruit and vegetables, dairy, red and processed meat, discretionary foods, and fluid intake. The specific foods contributing to each group are provided in Supplementary Table S1. Each component was scored from 0 to 5 points based on adherence to the Eatwell Guide recommendations, yielding a maximum total score of 60 points. Participants meeting the recommended intake for a component received 5 points. For foods considered beneficial (e.g., fruit and vegetables, wholegrains, fish), lower consumption received proportionally fewer points, with 0 points assigned for less than half the recommended intake. For foods considered less healthy (e.g., red and processed meat, discretionary foods), 0 points were awarded when consumption exceeded 1.5 times the recommended limit. Although dairy foods contribute important nutrients and form part of the core Eatwell Guide recommendations, overconsumption was scored as lower adherence, as high intakes may be associated with greater saturated fat intake.

### Outcome assessment

Markers of adiposity were assessed in line with UK Biobank protocols [19] at the baseline assessment centre and subsequent follow up visits. The primary outcome of BMI was calculated from weight and height measured by trained research staff using standardised procedures. BMI was used as a proxy for overall adiposity given its widespread quantification in the UK Biobank. Whilst BMI does not differentiate between fat mass and lean mass, or provide information on fat distribution, it provides a valid assessment of overall fat mass adjusted for height [25,26].

To address the limitations of BMI and provide a more nuanced assessment of adiposity, additional measures were included. To capture central fat accumulation, waist circumference and ABSI were measured. Central adiposity has been shown to be more strongly associated with cardiometabolic risk and mortality than BMI [16]. Waist circumference was measured as per UK Biobank protocols, at the midpoint between the lowest rib and the iliac crest using a non-stretchable tape. ABSI was calculated as waist circumference/BMI^2/3^ x height^1/2^ and a sex standardised z score was then calculated to control for sex differences [27]. Bioelectrical impedance analysis (BIA) (Tanita BC418MA) was used to provide valid [28], complementary measures of overall and regional fat distribution, including total body fat percentage and trunk fat percentage.

### Statistical analysis

All analyses were conducted in R Studio (version 4.4.0). Overall and stratified baseline characteristics of the analytic sample were summarised as mean ± SD for continuous variables and as percentages for categorical variables. To determine cross-sectional associations between Eatwell Guide adherence and markers of excess weight, linear regression analysis was conducted. Logistic regression analysis was conducted to determine associations between groups of Eatwell Guide adherence (low, medium and high) and markers of adiposity. In the prospective analysis, associations between Eatwell Guide (both continuously and categorically) and markers of adiposity were quantified over time via linear mixed model analysis. Analyses were adjusted simultaneously for: age, sex, ethnicity (white, other ethnic background), socio-economic status (Townsend Index categorised as low [quintile 1], moderate [quintiles 2–4], high [quintile 5] deprivation), education (higher [college/university/ other professional qualification], vocational [NVQ/HND/ HNC], upper secondary [A-levels], lower secondary [O-levels/GCSEs /CSEs] or none), smoking status (never, past, current), physical activity (international physical activity questionnaire [IPAQ] group, categorised as low, medium, high) and energy intake (kcal/d). For visualisation of model-predicted trajectories, predicted BMI values and 95% confidence intervals for each Eatwell Guide tertile (low, medium, high) across all timepoints were derived from the linear mixed-effects model, adjusting for all covariates.

Stratified analyses were conducted to determine whether associations between adherence to the Eatwell Guide and markers of adiposity were moderated by age (grouped as ‘younger’ vs. ‘older’ by dichotomising at the median (57 years)), sex (male vs. female), physical activity (low vs. moderate vs. high) and socioeconomic status (low vs. middle vs. most deprived). We also investigated whether associations between Eatwell Guide adherence and markers of adiposity were moderated by genetic risk of obesity (low vs. high risk). Genetic predisposition to adiposity was accounted for using a polygenic risk score for BMI [29]. This variable was derived from the genome-wide association study (GWAS) summary statistics from the GIANT consortium [30] and subsequently curated by UK Biobank.

### Sensitivity analysis

Sensitivity analyses tested the robustness of associations between Eatwell Guide adherence and adiposity markers. These included: restricting to participants with ≥2 dietary recalls; excluding extreme energy intakes (<800 or >4200 kcal/d for males; <600 or >3500 kcal/d for females); removing energy intake as a covariate; sequentially removing each Eatwell Guide component to assess whether any single component drove associations; and repeating genetic risk stratification using tertiles rather than dichotomous groups.

## Results

### Cohort characteristics

Of the 502 526 participants who underwent baseline assessment in the UK Biobank study, 156 764 were included in the present analysis. Baseline characteristics of the analytical cohort, overall and stratified by Eatwell Guide adherence level (low, medium and high) are provided in Table 1. Socio-demographic differences in Eatwell guide adherence in this cohort have been presented previously [24]. The analytical cohort was broadly similar to the remainder of the UK Biobank cohort, with minor differences in education and physical activity level (Supplementary Table 2).

**Table 1.**
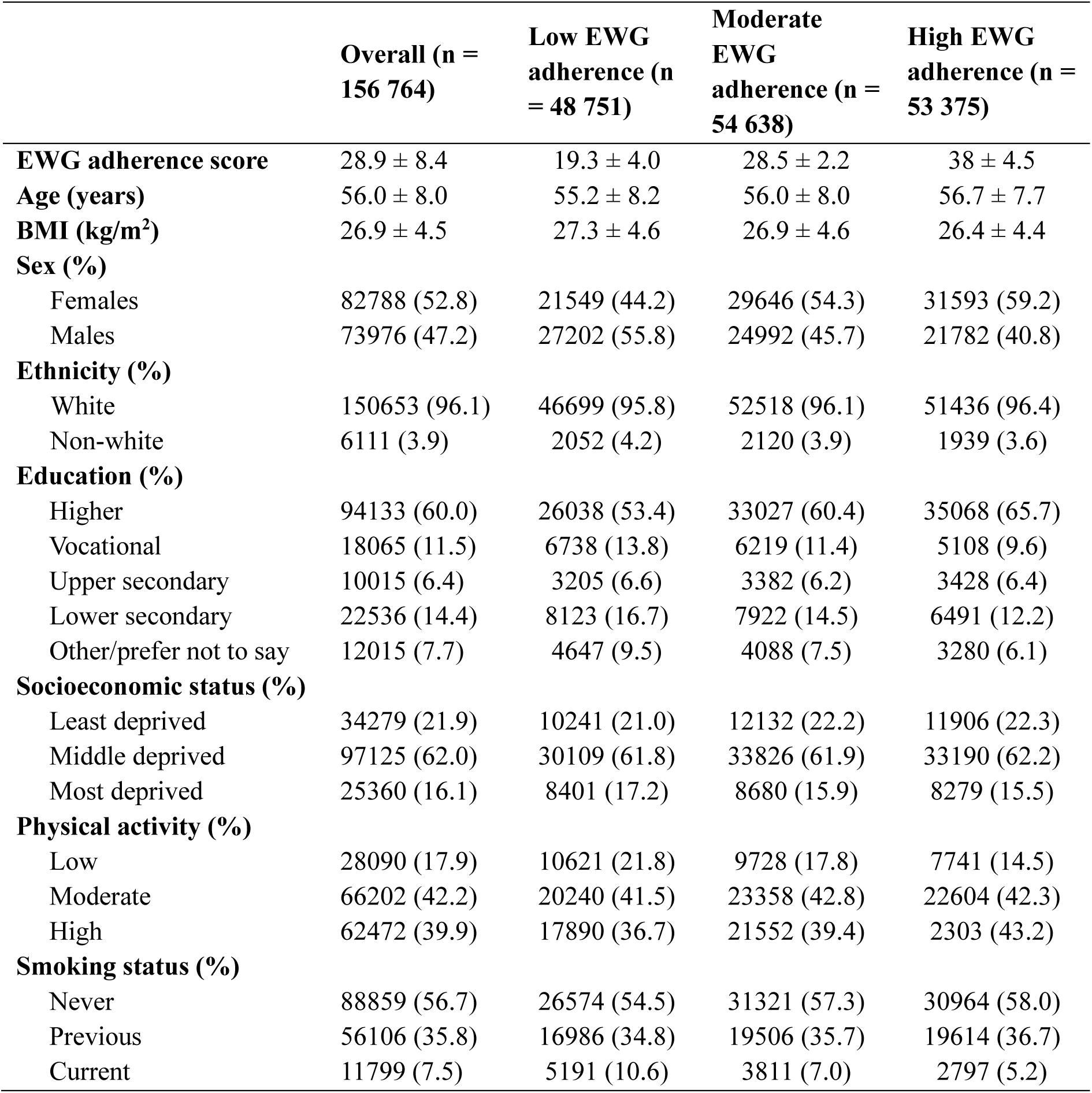
Baseline characteristics of analytical sample.

|  | **Overall (n =**<br>**156 764)** | **Low EWG**<br>**adherence (n**<br>**= 48 751)** | **Moderate**<br>**EWG**<br>**adherence (n =**<br>**54 638)** | **High EWG**<br>**adherence (n =**<br>**53 375)** |
| --- | --- | --- | --- | --- |
| **EWG adherence score** | 28.9 ± 8.4 | 19.3 ± 4.0 | 28.5 ± 2.2 | 38 ± 4.5 |
| **Age (years)** | 56.0 ± 8.0 | 55.2 ± 8.2 | 56.0 ± 8.0 | 56.7 ± 7.7 |
| **BMI (kg/m<sup>2</sup>)** | 26.9 ± 4.5 | 27.3 ± 4.6 | 26.9 ± 4.6 | 26.4 ± 4.4 |
| **Sex (%)** |  |  |  |  |
| Females | 82788 (52.8) | 21549 (44.2) | 29646 (54.3) | 31593 (59.2) |
| Males | 73976 (47.2) | 27202 (55.8) | 24992 (45.7) | 21782 (40.8) |
| **Ethnicity (%)** |  |  |  |  |
| White | 150653 (96.1) | 46699 (95.8) | 52518 (96.1) | 51436 (96.4) |
| Non-white | 6111 (3.9) | 2052 (4.2) | 2120 (3.9) | 1939 (3.6) |
| **Education (%)** |  |  |  |  |
| Higher | 94133 (60.0) | 26038 (53.4) | 33027 (60.4) | 35068 (65.7) |
| Vocational | 18065 (11.5) | 6738 (13.8) | 6219 (11.4) | 5108 (9.6) |
| Upper secondary | 10015 (6.4) | 3205 (6.6) | 3382 (6.2) | 3428 (6.4) |
| Lower secondary | 22536 (14.4) | 8123 (16.7) | 7922 (14.5) | 6491 (12.2) |
| Other/prefer not to say | 12015 (7.7) | 4647 (9.5) | 4088 (7.5) | 3280 (6.1) |
| **Socioeconomic status (%)** |  |  |  |  |
| Least deprived | 34279 (21.9) | 10241 (21.0) | 12132 (22.2) | 11906 (22.3) |
| Middle deprived | 97125 (62.0) | 30109 (61.8) | 33826 (61.9) | 33190 (62.2) |
| Most deprived | 25360 (16.1) | 8401 (17.2) | 8680 (15.9) | 8279 (15.5) |
| **Physical activity (%)** |  |  |  |  |
| Low | 28090 (17.9) | 10621 (21.8) | 9728 (17.8) | 7741 (14.5) |
| Moderate | 66202 (42.2) | 20240 (41.5) | 23358 (42.8) | 22604 (42.3) |
| High | 62472 (39.9) | 17890 (36.7) | 21552 (39.4) | 2303 (43.2) |
| **Smoking status (%)** |  |  |  |  |
| Never | 88859 (56.7) | 26574 (54.5) | 31321 (57.3) | 30964 (58.0) |
| Previous | 56106 (35.8) | 16986 (34.8) | 19506 (35.7) | 19614 (36.7) |
| Current | 11799 (7.5) | 5191 (10.6) | 3811 (7.0) | 2797 (5.2) |

### Cross-sectional associations between Eatwell Guide adherence and markers of adiposity

In the cross-sectional analyses using linear regression, higher adherence to the Eatwell Guide was associated with lower markers of adiposity, including BMI, waist circumference, ABSI and both total and trunk-specific body fat (all p<0.001, Supplementary Table S3).

Similarly, in logistic regression analyses, both moderate and high Eatwell Guide adherence were associated with lower odds of all adiposity-related outcomes relative to low adherence (Supplementary Table S4). Moderate and high adherence were associated with 9% and 25% lower odds of overweight or obesity, 9–24% lower odds of elevated waist circumference, 10–15% lower odds of higher ABSI, 8–22% lower odds of higher body fat percentage, and 10–22% lower odds of higher trunk fat percentage (all p<0.001).

### Prospective associations between Eatwell Guide adherence and markers of adiposity

In the prospective linear mixed model analysis, higher Eatwell Guide adherence over time was associated with lower markers of adiposity, including BMI, waist circumference, total body fat percentage and trunk-specific body fat (all p<0.001, Table 2), with a trend observed for ABSI (*p* = 0.06).

**Table 2.** Linear mixed model analysis of prospective association with Eatwell Guide adherence score and markers of weight.

| Predictor (EWG score x time) | Beta | SE | P-value |
| --- | --- | --- | --- |
| BMI (n = 156 898) | -0.0080 | 0.00063 | <0.001 |
| Waist circumference (n = 157 051) | -0.0161 | 0.0023 | <0.001 |
| ABSI (n = 156 834) | 0.00056 | 0.00029 | 0.06 |
| Total body fat % (155 236) | -0.0151 | 0.00124 | <0.001 |
| Trunk body fat % (155 239) | -0.0171 | 0.00151 | <0.001 |

In the linear mixed model analysis of Eatwell Guide adherence split by tertiles, both moderate and higher adherence were associated with more favourable adiposity trajectories over time compared to low adherence (Table 3). A dose-response pattern was evident, with high adherence showing stronger associations than moderate adherence for BMI, waist circumference and total body fat percentage (all *p* < 0.001). For trunk body fat percentage, moderate and high adherence showed similar associations (both p<0.001). No significant associations were observed between adherence tertiles and ABSI trajectories (moderate: p=0.21; high: p=0.06).

**Table 3.** Linear mixed model analysis of prospective associations between Eatwell Guide adherence tertile and markers of adiposity.

| Predictor (EWG tertile x time) | Beta | SE | P-value |
| --- | --- | --- | --- |
| BMI (n = 156 898) |  |  |  |
| Low (ref) | - | - | - |
| Medium | -0.080 | 0.013 | <0.001 |
| High | -0.149 | 0.013 | <0.001 |
| Waist circumference (n = 157 051) |  |  |  |
| Low (ref) | - | - | - |
| Medium | -0.193 | 0.047 | <0.001 |
| High | -0.291 | 0.047 | <0.001 |
| ABSI (n = 156 834) |  |  |  |
| Low (ref) | - | - | - |
| Medium | 0.008 | 0.006 | 0.21 |
| High | 0.012 | 0.006 | 0.06 |
| Total body fat % (n = 155 236) |  |  |  |
| Low (ref) | - | - | - |
| Medium | -0.120 | 0.026 | <0.001 |
| High | -0.281 | 0.026 | <0.001 |
| Trunk body fat % (n = 155 239) |  |  |  |
| Low (ref) | - | - | - |
| Medium | -0.134 | 0.032 | <0.001 |
| High | -0.138 | 0.031 | <0.001 |

Trajectories of adiposity markers over time by Eatwell Guide adherence tertile are shown in Figure 1. The diverging trajectories demonstrate that higher adherence was associated with more favourable outcomes, except for ABSI which was not significantly different between tertiles.

**Figure 1.**
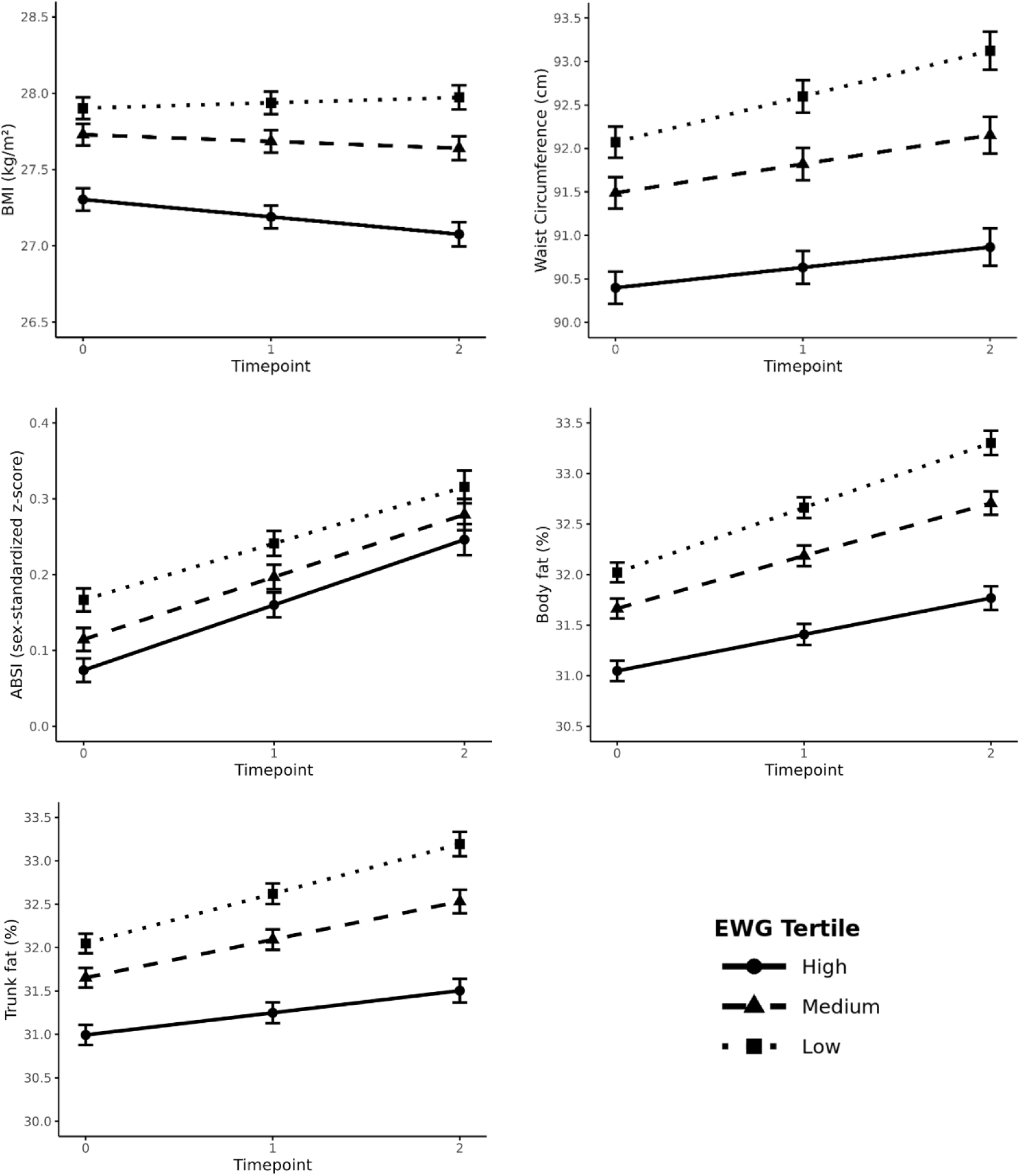
Predicted trajectories of BMI, waist circumference, ABSI z-score, total body fat percentage, and trunk fat percentage across EWG adherence tertiles over time. Models were adjusted for baseline age, sex, deprivation, education level, smoking status, physical activity, energy intake and ethnicity.

### Sub-group analyses

Sub-group analyses are presented in Supplementary Tables S5-9. Across all sub-groups examined, adherence to the Eatwell Guide was consistently associated with more favourable adiposity outcomes. Significant inverse associations with BMI, waist circumference, body fat percentage, and trunk fat percentage were observed regardless of sex, age, physical activity, socioeconomic status, ethnicity or genetic risk.

By sex, differences were minimal, although stronger effects were seen in females for BMI, both in the high adherence group (*p* = 0.04) and in the continuous model (*p* = 0.02). By age, effects were generally stronger in younger individuals, particularly for BMI and waist circumference, with significant interactions in the continuous models (p < 0.001 and p < 0.01, respectively). By socioeconomic status, stronger effects were noted among participants in the most deprived group, particularly for BMI and waist circumference, with evidence of interaction in both the high adherence categorical (p = 0.02) and continuous models (p = 0.04). There was no evidence of interaction effects by physical activity (all p > 0.40) or polygenic risk score (all p > 0.20). The non-significant effect of polygenic risk score remained when split by tertiles as a sensitivity analysis (Supplementary Table S10).

### Sensitivity analysis

The associations between higher Eatwell Guide adherence and more favourable trajectories of markers of adiposity over time were consistent to a range of sensitivity analyses. Results were similar when we repeated analyses for participants with a minimum of two dietary recalls (Supplementary Table S11), excluding participants with extreme energy intakes (Supplementary Table S12) and removing energy intake as a covariate from the model (supplementary Table S13). Prospective associations with Eatwell Guide adherence score and ABSI became significant (*p* = 0.04) following removal of extreme energy intakes. After sequential removal of individual dietary components, the associations between Eatwell Guide adherence and BMI, waist circumference, body fat percentage and trunk fat percentage remained consistent (Supplementary Tables S14, S15, S17 and 18). When white meat (p = 0.02), dairy (p = 0.048), beans and pulses (p = 0.02), nuts (p = 0.047), eggs (p = 0.02), and discretionary foods (p = 0.04) were removed from the score, the associations between Eatwell Guide adherence and ABSI became significant, indicating a less favourable relationship (Supplementary Table S16).

## Discussion

This study represents the first large-scale prospective analysis examining associations between Eatwell Guide adherence and multiple markers of adiposity. Higher Eatwell Guide adherence was inversely associated with multiple markers of adiposity both cross-sectionally and prospectively in the UK Biobank cohort. A dose-response relationship was observed for BMI, waist circumference, total body fat percentage and trunk fat percentage, with inconsistent associations for ABSI, likely a result of capturing body shape as opposed to adiposity. Similar patterns were observed across multiple population subgroups, including those with different levels of genetic risk for obesity. This suggests that greater adherence to the Eatwell Guide is associated with benefits irrespective of genetic predisposition or demographic characteristics.

Cross-sectional data from the present study demonstrates 25% lower odds of overweight or obesity with higher adherence to the Eatwell Guide. This is supported by our prospective analysis, which suggests sustained benefits over time in a dose response manner. These findings are consistent with previous cross-sectional work [13], but in contrast to the only previous prospective study conducted in post-menopausal women [14], likely explained by the larger sample size and stronger statistical power in the present study. Interestingly, when energy intake was removed from the models, associations remained similar, suggesting that energy intake differences contributed minimally to the observed relationships. These findings may be explained by differences in food matrix and/or level of processing, which may influence energy bioavailability. Foods that align with the Eatwell Guide tend to have complex food matrices rich in dietary fibre, reducing the metabolisable energy derived from these foods [31,32]. Recent evidence supports the importance of food processing status within the Eatwell Guide. Dicken et al., [33] found that adults following an Eatwell Guide diet composed of minimally processed foods lost significantly more weight than those consuming ultra-processed foods, likely due to differences in palatability and energy bioavailability via food matrix alterations [34].

Our findings related to fat distribution build upon Best & Flannery’s [14] observations of prospective reductions in waist circumference by conducting a more comprehensive analysis of fat distribution and body composition. We observed reductions in waist circumference but also demonstrated inverse associations between Eatwell Guide adherence and trunk and total body fat percentage. This suggests that the benefits of adhering to the Eatwell Guide are not regionally specific but reflect whole body improvements in body composition. Improvements in trunk fat percentage are particularly noteworthy given that visceral and subcutaneous abdominal fat are associated with insulin resistance, dyslipidemia, and systemic inflammation [35]. Importantly, these findings demonstrate that the benefits of Eatwell Guide adherence are not merely related to weight loss but translate to beneficial changes in body composition. Of particular relevance, pharmacological interventions for obesity treatment such as glucagon-like peptide-1 receptor agonists, have shown less favourable changes in body composition (including loss of muscle mass), which may have implications for longer term metabolic health [36]. Data from this study support the need for better dietary support alongside and after pharmacological treatment, to ensure optimal improvements in body composition [37].

Subgroup analyses demonstrated that associations were broadly consistent across demographic groups, suggesting universal benefits of Eatwell Guide adherence. Of note, there was no evidence of effect modification by physical activity level, highlighting diet quality as an independent determinant of body composition and weight. Whilst physical activity induces substantial cardiovascular and metabolic health benefits [38], this finding highlights the role of dietary quality as the primary factor in the management of weight and adiposity. It is however worth emphasising that a comprehensive healthy lifestyle approach incorporating both diet and physical activity remains essential for overall health [39]. The lack of moderation by polygenic risk score indicates that individuals genetically predisposed to obesity derive similar benefits from adhering to the Eatwell Guide as those at lower genetic risk. Despite this, it is acknowledged that adhering to healthy dietary patterns may present differential challenges, potentially a result of biological factors such as dysregulated appetite, or other social, cultural, emotional or environmental determinants [40]. As such, whilst the Eatwell Guide provides an effective dietary pattern in the context of weight and adiposity, implementation of such dietary recommendations should employ person-centred approaches that recognise and address these multifactorial barriers to dietary adherence and ensure equitable access [5].

This study has several strengths. Firstly, our study had a large sample size (n=156,764), building on previous studies that evaluated Eatwell Guide adherence in smaller, more specialised cohorts. This was also the first study to examine how genetic predisposition to obesity may influence the association between Eatwell Guide intake and adiposity, using a comprehensive polygenic risk score. Additionally, we assessed multiple markers of adiposity, providing the first comprehensive overview of the relationship between Eatwell Guide and body composition, and employed a prospective study design which allowed us to explore adiposity trajectories over time. The use of a food-based scoring system which closely aligns with public health messaging also maximises the practical relevance of our findings for policy and practice. Finally, we conducted a range of sensitivity analyses which our data remained robust to, such as only including participants with a minimum of two dietary reports, excluding individuals with extreme energy intakes, and sequentially removing a component of the Eatwell Guide in turn to see if any particular component drove the associations.

Several limitations should also be acknowledged. First, the observational design of the present study limits causal inference, and it is possible the findings may be a result of reverse causality. Nevertheless, our prospective design and exploration of adiposity trajectories help mitigate this risk. Second, self-reported dietary intake may be subject to measurement error, social desirability bias, and potential underreporting, particularly among individuals with overweight or obesity [41]. In addition, some participants contributed only one or two assessments, which may not fully represent usual dietary patterns, however previous work in UK Biobank has demonstrated stability of dietary reporting over time [42]. Third, the UK Biobank cohort is susceptible to healthy volunteer bias, with individuals also generally more affluent and more highly educated than the UK general population [43], which may may somewhat limit the generalisability of the findings. Fourth, the predominantly white (96%) analytical sample restricted subgroup analyses by ethnicity, a key demographic consideration given the diversity of cultural dietary patterns, varying obesity prevalence and adiposity deposition across ethnic groups in the UK [44]. Fifth, residual confounding remains a potential limitation of this study, despite covariate adjustment. Sixth, more recent and larger GWAS for BMI exist [45], however the polygenic risk score used here has been extensively validated [46]. Finally, the translation of qualitative dietary guidance for some components (e.g. eat more beans and pulses), into quantitative recommendations required some interpretative decisions to create the scoring system.

The policy and practice implications considered within this study are summarised in Figure 2. Future research is required to establish causality between Eatwell Guide adherence and markers of adiposity using randomised controlled trials. Future research should also be conducted using cohorts with a greater representation of ethnic minorities, using culturally appropriate Eatwell Guide scores derived from adapted Eatwell Guide models [47,48]. There is also a need to explore barriers and facilitators to Eatwell Guide adherence to inform the development of relevant behaviour change interventions to improve intake across the UK.

**Figure 2.**
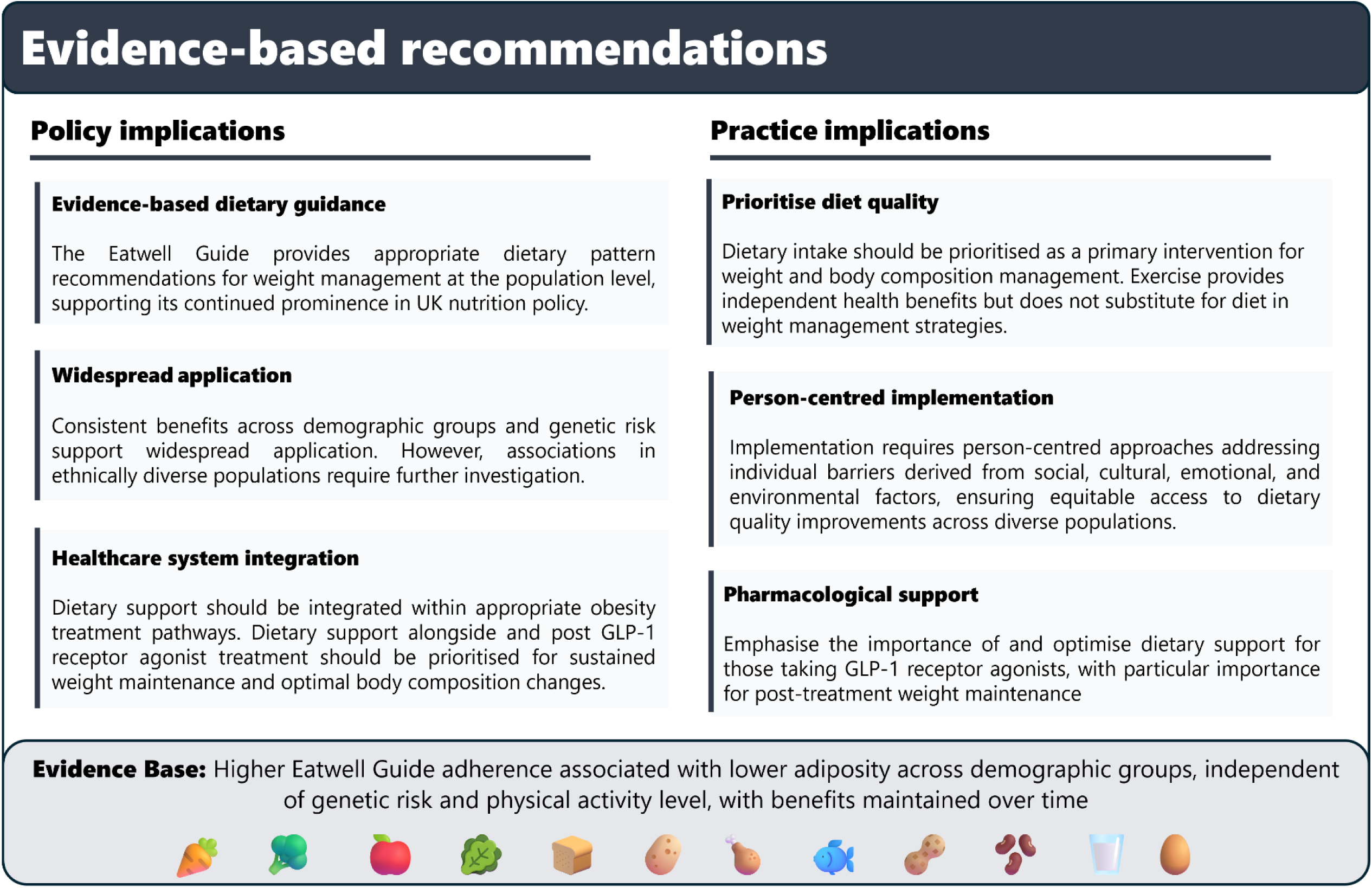
Summary of the policy and practice recommendations

In conclusion, this large-scale prospective cohort study demonstrated that adherence to the UK healthy eating model (the Eatwell Guide) was associated with beneficial changes in multiple markers of adiposity over time. These effects were consistent across several demographic groups and irrespective of physical activity level and genetic risk of obesity. These findings establish the importance of diet quality in weight management but highlight the need for further research to explore appropriate behaviour change interventions to improve adherence in the UK population.

## Data Availability

The data that support the findings of this study are available from UK Biobank, but restrictions apply to the availability of these data, which were used under license for this study, and so are not publicly available. Data are however available from the authors upon reasonable request and with permission of UK Biobank.

## Acknowledgements

This research has been conducted using the UK Biobank Resource under application number 147784.

## Author Contributions

AG and OS conceived the idea and generated the aim of the research. AG, JM, SG, and OS developed the adherence tool and AG, SG and OS conducted statistical analysis. All authors reviewed findings, contributed to the discussion and provided review of manuscript. No funding was received for this project.

## Competing Interests

The authors declare no competing interests.

## Notes

### Competing Interest Statement

The authors have declared no competing interest.

### Funding Statement

The study did not receive any funding

### Author Declarations

Ethical approval for the UK Biobank study was provided by the North West Haydock Research Ethics Committee (REC reference: 16/NW/0274), and all participants provided electronic signed consent.

## References

1. NHS Digital. Health Survey for England, 2022 Part 2. 2024.

2. Koliaki C, Liatis S, Kokkinos A. Obesity and cardiovascular disease: revisiting an old relationship. Metabolism. 2019;92:98–107.

3. Gunter MJ, Berrington De Gonzalez A. Is obesity a cause of all cancer types? The Lancet Regional Health - Europe. 2024;46:101110.

4. Hirko K, Kantor E, Cohen S, Blot W, Stampfer M, Signorello L. Body mass index in young adulthood, obesity trajectory, and premature mortality. American Journal of Epidemiology. 2015;182(5):441–50.

5. Ells L, Ashton M, Li R, Logue J, Griffiths C, Torbahn G, et al. Can we deliver person-centred obesity care across the globe? Current Obesity Reports. 2022;11(4):350–5.

6. Torbahn G, Jones A, Griffiths A, Matu J, Metzendorf M, Ells LJ, et al. Pharmacological interventions for the management of children and adolescents living with obesity—An update of a Cochrane systematic review with meta-analyses. Pediatric Obesity. 2024;19(5):e13113.

7. Yao H, Zhang A, Li D, Wu Y, Wang CZ, Wan JY, et al. Comparative effectiveness of GLP-1 receptor agonists on glycaemic control, body weight, and lipid profile for type 2 diabetes: systematic review and network meta-analysis. BMJ. 2024;e076410.

8. Fallows E, Ells L, Anand V. Semaglutide and the future of obesity care in the UK. The Lancet. 2023;401(10394):2093–6.

9. PHE. The Eatwell Guide. 2016; Available from: https://assets.publishing.service.gov.uk/media/5a75564fed915d6faf2b2375/Eatwell_guide_colour.pdf

10. Scarborough P, Kaur A, Cobiac L, Owens P, Parlesak A, Sweeney K, et al. Eatwell Guide: modelling the dietary and cost implications of incorporating new sugar and fibre guidelines. BMJ Open. 2016;6:13182–13182.

11. Scheelbeek P, Green R, Papier K, Knuppel A, Alae-Carew C, Balkwill A, et al. Health impacts and environmental footprints of diets that meet the Eatwell Guide recommendations: analyses of multiple UK studies. BMJ Open. 2020;10:37554–37554.

12. Fadnes L, Celis-Morales C, Økland J, Parra-Soto S, Livingstone K, Ho F, et al. Life expectancy can increase by up to 10 years following sustained shifts towards healthier diets in the United Kingdom. Nature Food. 2023;4:961–5.

13. Gregory S, Griffiths A, Jennings A, Malcomson F, Matu J, Minihane A, et al. Adherence to the Eatwell Guide and cardiometabolic, cognitive and neuroimaging parameters: an analysis from the PREVENT dementia study. Nutrition and Metabolism. 2024;21(1):21–21.

14. Best N, Flannery O. Association between adherence to the Mediterranean Diet and the Eatwell Guide and changes in weight and waist circumference in post-menopausal women in the UK Women’s Cohort Study. Post Reproductive Health. 2023;29(1):25–32.

15. Nuttall FQ. Body Mass Index: Obesity, BMI, and Health A Critical Review. Nutrition Today. 2015;50(3):117–28.

16. Franek E, Pais P, Basile J, Nicolay C, Raha S, Hickey A, et al. General versus central adiposity as risk factors for cardiovascular-related outcomes in a high-risk population with type 2 diabetes: a post hoc analysis of the REWIND trial. Cardiovasc Diabetol. 2023;22(1):52.

17. Kim MS, Shim I, Fahed AC, Do R, Park WY, Natarajan P, et al. Association of genetic risk, lifestyle, and their interaction with obesity and obesity-related morbidities. Cell Metabolism. 2024;36(7):1494–1503.e3.

18. Ollier W, Sprosen T, Peakman T. UK Biobank: from concept to reality. Pharmacogenomics. 2005;6:639–46.

19. Sudlow C, Gallacher J, Allen N, Beral V, Burton P, Danesh J, et al. UK Biobank: An Open Access Resource for Identifying the Causes of a Wide Range of Complex Diseases of Middle and Old Age. PLOS Medicine. 2015;e1001779.

20. Schorr M, Miller KK. The endocrine manifestations of anorexia nervosa: mechanisms and management. Nat Rev Endocrinol. 2017;13(3):174–86.

21. Setiawan T, Sari IN, Wijaya YT, Julianto NM, Muhammad JA, Lee H, et al. Cancer cachexia: molecular mechanisms and treatment strategies. J Hematol Oncol. 2023;16(1):54.

22. Greenwood D, Hardie L, Frost G, Alwan N, Bradbury K, Carter M, et al. Validation of the Oxford WebQ Online 24-Hour Dietary Questionnaire Using Biomarkers. American Journal of Epidemiology. 2019;188:1858–67.

23. Liu B, Young H, Crowe F, Benson V, Spencer E, Key T, et al. Development and evaluation of the Oxford WebQ, a low-cost, web-based method for assessment of previous 24 h dietary intakes in large-scale prospective studies. Public Health Nutrition. 2011;14:1998–2005.

24. Griffiths A, Malcomson F, Matu J, Gregory S, Fairley AM, Townsend RF, et al. Socio-demographic variation in adherence to The Eatwell Guide within the UK Biobank prospective cohort study [Internet]. Nutrition; 2025 [cited 2025 Oct 15]. Available from: http://medrxiv.org/lookup/doi/10.1101/2025.06.06.25329110

25. Willett W. Nutritional Epidemiology. 3rd edn. Oxford University Press; 2013.

26. Spiegelman D, Israel R, Bouchard C, Willett W. Absolute fat mass, percent body fat, and body-fat distribution: which is the real determinant of blood pressure and serum glucose? The American Journal of Clinical Nutrition. 1992;55(6):1033–44.

27. Krakauer NY, Krakauer JC. A New Body Shape Index Predicts Mortality Hazard Independently of Body Mass Index. Li S, editor. PLoS ONE. 2012;7(7):e39504.

28. Feng Q, Bešević J, Conroy M, Omiyale W, Lacey B, Allen N. Comparison of body composition measures assessed by bioelectrical impedance analysis versus dual-energy X-ray absorptiometry in the United Kingdom Biobank. Clinical Nutrition ESPEN. 2024;63:214–25.

29. Thompson DJ, Wells D, Selzam S, Peneva I, Moore R, Sharp K, et al. A systematic evaluation of the performance and properties of the UK Biobank Polygenic Risk Score (PRS) Release. Zhu X, editor. PLoS ONE. 2024;19(9):e0307270.

30. Locke AE, Kahali B, Berndt SI, Justice AE, Pers TH, Day FR, et al. Genetic studies of body mass index yield new insights for obesity biology. Nature. 2015;518(7538):197–206.

31. Baer DJ, Rumpler WV, Miles CW, Fahey GC. Dietary Fiber Decreases the Metabolizable Energy Content and Nutrient Digestibility of Mixed Diets Fed to Humans. The Journal of Nutrition. 1997;127(4):579–86.

32. Capuano E, Oliviero T, Fogliano V, Pellegrini N. Role of the food matrix and digestion on calculation of the actual energy content of food. Nutrition Reviews. 2018;76(4):274–89.

33. Dicken SJ, Jassil FC, Brown A, Kalis M, Stanley C, Ranson C, et al. Ultraprocessed or minimally processed diets following healthy dietary guidelines on weight and cardiometabolic health: a randomized, crossover trial. Nat Med [Internet]. 2025 [cited 2025 Oct 15]; Available from: https://www.nature.com/articles/s41591-025-03842-0

34. Forde CG, Bolhuis D. Interrelations Between Food Form, Texture, and Matrix Influence Energy Intake and Metabolic Responses. Curr Nutr Rep. 2022;11(2):124–32.

35. Hardy OT, Czech MP, Corvera S. What causes the insulin resistance underlying obesity? Current Opinion in Endocrinology, Diabetes & Obesity. 2012;19(2):81–7.

36. Prado CM, Phillips SM, Gonzalez MC, Heymsfield SB. Muscle matters: the effects of medically induced weight loss on skeletal muscle. The Lancet Diabetes & Endocrinology. 2024;12(11):785–7.

37. Spreckley M, Ruggiero CF, Brown A. Bridging the nutrition guidance gap for GLP-1 receptor agonist therapy assisted weight loss: lessons from bariatric surgery. Int J Obes [Internet]. 2025 [cited 2025 Nov 26]; Available from: https://www.nature.com/articles/s41366-025-01952-w

38. Nystoriak MA, Bhatnagar A. Cardiovascular Effects and Benefits of Exercise. Front Cardiovasc Med. 2018;5:135.

39. Moradell A, Casajús JA, Moreno LA, Vicente-Rodríguez G, Gómez-Cabello A. Effects of Diet-Exercise Interaction on Human Health across a Lifespan. Nutrients. 2023;15(11):2520.

40. Deslippe AL, Soanes A, Bouchaud CC, Beckenstein H, Slim M, Plourde H, et al. Barriers and facilitators to diet, physical activity and lifestyle behavior intervention adherence: a qualitative systematic review of the literature. Int J Behav Nutr Phys Act. 2023;20(1):14.

41. Hebert JR, Clemow L, Pbert L, Ockene IS, Ockene JK. Social Desirability Bias in Dietary Self-Report May Compromise the Validity of Dietary Intake Measures. Int J Epidemiol. 1995;24(2):389–98.

42. Bradbury KE, Young HJ, Guo W, Key TJ. Dietary assessment in UK Biobank: an evaluation of the performance of the touchscreen dietary questionnaire. J Nutr Sci. 2018;7:e6.

43. Fry A, Littlejohns T, Sudlow C, Doherty N, Adamska L, Sprosen T, et al. Comparison of sociodemographic and health-related characteristics of UK Biobank participants with those of the general population. American Journal of Epidemiology. 2017;186(9):1026–34.

44. Ojo AS, Nnyanzi LA, Giles EL, Ells L, Okeke SR, Ajayi KV, et al. “I am not really into the government telling me what I need to eat”: exploring dietary beliefs, knowledge, and practices among ethnically diverse communities in England. BMC Public Health. 2023;23(1):800.

45. Yengo L, Sidorenko J, Kemper KE, Zheng Z, Wood AR, Weedon MN, et al. Meta-analysis of genome-wide association studies for height and body mass index in ∼700000 individuals of European ancestry. Human Molecular Genetics. 2018;27(20):3641–9.

46. Khera AV, Chaffin M, Wade KH, Zahid S, Brancale J, Xia R, et al. Polygenic Prediction of Weight and Obesity Trajectories from Birth to Adulthood. Cell. 2019;177(3):587–596.e9.

47. Jay F. South Asian Eatwell Guide. South Asian Eatwell Guide [Internet]. 2021; Available from: https://mynutriweb.com/wp-content/uploads/2021/10/Untitled-700-x-700-px.pdf

48. Saint Hill A. The African and Caribbean Eatwell Guide. The African and Caribbean Eatwell Guide [Internet]. 2023; Available from: https://mynutriweb.com/the-african-and-caribbean-eatwell-guide/

