## Supplementary materials for "Adherence to the Eatwell Guide and associations with markers of adiposity: A prospective analysis within the UK Biobank cohort"

**Supplementary information**

**Supplementary Table S1 –** List of individual foods which contributed to each food group and the Eatwell Guide scoring methodology

| Food component | Contributing foods from the Oxford WebQ | Average consumption required (servings/day) | EWG Score |
|---|---|---|---|
| **Fruit and vegetables** | Stewed fruit (104410), Prune (104420), Dried fruit (104430), Mixed fruit (104440), Apple (104450), Banana (104460), Berry (104470), Cherry (104480), Grapefruit (104490), Grape (104500), Mango (104510), Melon (104520), Orange (104530), Satsuma (104540), Peach nectarine (104550), Pear (104560), Pineapple (104570), Plum (104580), Other fruit (104590), Orange juice (100190), Grapefruit juice (100200), Pure fruit vegetable juice (100210), Fruit smoothie (100220), Mixed veg (104060), Veg pieces (104070), Coleslaw (104080), Side salad (104090), Avocado (104100), Beetroot (104130), Broccoli (104140), Butternut squash (104150), Cabbage kale (104160), Carrot (104170), Cauliflower (104180), Celery (104190), Courgette (104200), Cucumber (104210), Garlic (104220), Leek (104230), Lettuce (104240), Mushroom (104250), Onion (104260), Parsnip (104270), Sweet pepper (104290), Spinach (104300), Sprouts (104310), Sweetcorn (104320), Fresh tomato (104340), Tinned tomato (104350), Turnip swede (104360), Watercress (104370), Other veg (104380), Olive (102490) | From 0 to < 2.5 | 0 |
| | | From 2.5 to < 3.125 | 1 |
| | | From 3.125 to < 3.75 | 2 |
| | | From 3.75 to < 4.375 | 3 |
| | | From 4.375 to < 5 | 4 |
| | | ≥5 | 5 |
| **Starchy Carbohydrates** | White pasta (102710), Wholemeal pasta (102720), White rice (102730), Brown rice (102740), Snackpot (102760), Couscous (102770), Other grain (102780), Sliced bread (100950), Baguette (101020), Bap (101090), Bread roll (101160), Naan bread (101230), Garlic bread (101240), Crispbread (101250), Oatcake (101260), Other bread (101270), Porridge (100770), Muesli (100800), Oat crunch (100810), Plain cereal (100830), Bran cereal (100840), Wholewheat cereal (100850), Other cereal (100860), Fried potatoes (104020), Boiled baked potatoes (104030), Mashed potato (104050), Sweet potato (104330) | **Men**<br>< 2.5<br>**Women**<br>< 2 | 0 |
| | | **Men**<br>From 2.5 to < 3.125<br>**Women** | 1 |

| | | | |
|---|---|---|---|
| | | From 2 to < 2.5 | |
| | | **Men**<br>From 3.125 to < 3.75<br>**Women**<br>From 2.5 to < 3 | 2 |
| | | **Men**<br>From 3.75 to < 4.375<br>**Women**<br>From 3 to < 3.5 | 3 |
| | | **Men**<br>From 4.375 to < 5<br>**Women**<br>From 3.5 to < 4 | 4 |
| | | **Men**<br>≥ 5<br>**Women**<br>≥ 4 | 5 |
| **Wholegrains** | Wholemeal pasta (102720), Brown rice (102740), Sliced bread (100950), Type of bread (20091), Baguette (101020), Type of baguette (20092), Bap (101090), Type of bap (20093), Bread roll (101160), Type of bread roll (20094), Oatcake (101260), Porridge | From 0 to < 1.5 | 0 |
| | | From 1.5 to < 1.875 | 1 |
| | | From 1.875 to < 2.25 | 2 |

| | | | |
|---|---|---|---|
| | (100770), Muesli (100800), Bran cereal (100840), Wholewheat cereal (100850), Other grain (102780) | From 2.25 to < 2.625 | 3 |
| | | From 2.625 to < 3 | 4 |
| | | ≥ 3 | 5 |
| **Beans/pulses** | Pea (104280), Green beans (104120), Broad beans (104110), Baked beans (104000), Pulses (104010), Tofu (103270) | From 0 to < 0.21 | 0 |
| | | From 0.21 to < 0.27 | 1 |
| | | From 0.27 to < 0.32 | 2 |
| | | From 0.32 to < 0.38 | 3 |
| | | From 0.38 to < 0.43 | 4 |
| | | ≥ 0.43 | 5 |
| **Fish**[a] | Tinned tuna (103150), Oily fish (103160), Breaded fish (103170), Battered fish (103180), White fish (103190), Prawns (103200), Lobster crab (103210), Shellfish (103220), Other fish (103230) | **Fish**<br>From 0 to < 0.14<br>**Oily fish**<br>From 0 to < 0.07 | 0 |
| | | **Fish**<br>From 0.14 to < 0.18<br>**Oily fish**<br>From 0.07 to < 0.09 | 1 |
| | | **Fish**<br>From 0.18 to < 0.21<br>**Oily fish** | 2 |

| | | | |
|---|---|---|---|
| | | From 0.09 to < 0.11 | |
| | | **Fish**<br>From 0.21 to < 0.25<br>**Oily fish**<br>From 0.11 to < 0.13 | 3 |
| | | **Fish**<br>From 0.25 to < 0.29<br>**Oily fish**<br>From 0.13 to < 0.14 | 4 |
| | | **Fish**<br>≥ 0.29<br>**Oily fish**<br>≥ 0.14 | 5 |
| **Poultry** | Poultry (103060), Breaded poultry (103050) | From 0 to < 0.07 | 0 |
| | | From 0.07 to < 0.09 | 1 |
| | | From 0.09 to < 0.11 | 2 |
| | | From 0.11 to < 0.13 | 3 |
| | | From 0.13 to < 0.14 | 4 |
| | | ≥ 0.14 | 5 |
| **Nuts** | | From 0 to < 0.07 | 0 |

| | | | |
|---|---|---|---|
| | Unsalted nuts (102440), salted nuts (102430), unsalted peanuts (102420), salted peanuts (102410) | From 0.07 to < 0.09 | 1 |
| | | From 0.09 to < 0.11 | 2 |
| | | From 0.11 to < 0.13 | 3 |
| | | From 0.13 to < 0.14 | 4 |
| | | ≥ 0.14 | 5 |
| **Eggs** | Whole egg (102940), Omelette (102950), Egg sandwiches (102960), Scotch egg (102970), Other egg (102980) | From 0 to < 0.07 | 0 |
| | | From 0.07 to < 0.09 | 1 |
| | | From 0.09 to < 0.11 | 2 |
| | | From 0.11 to < 0.13 | 3 |
| | | From 0.13 to < 0.14 | 4 |
| | | ≥ 0.14 | 5 |
| **Red and processed meat** | Bacon (103070), Ham (103080), Liver (103090), Sausage (103010), Beef (103020), Pork (103030), Lamb (103040) | ≥ 1.5 | 0 |
| | | From 1.375 to < 1.5 | 1 |
| | | From 1.25 to < 1.375 | 2 |
| | | From 1.12 To < 1.25 | 3 |
| | | From 1 to < 1.12 | 4 |
| | | < 1 | 5 |
| **Dairy** | Yogurt (102090), Low fat hard cheese (102810), Hard cheese (102820), Low fat cheese spread (102850), Cheese spread (102860), Soft cheese (102830), Goat cheese (102900), Blue cheese (102840), Feta (102880), Mozzarella (102890), Other cheese (102910), Cottage cheese (102870), Milk (100520), Flavoured milk (100530), Added milk instant | ≥ 3 | 0 |
| | | From 2.75to < 3 | 1 |
| | | From 2.5 to < 2.75 | 2 |

| | | | |
|---|---|---|---|
| | coffee (100260), Added milk filtered coffee (100280), Added milk espresso (100320), Added milk other coffee (100350), Added milk standard tea (100460), Added milk rooibos tea (100480), Instant coffee (100250), Filtered coffee (100270), Espresso (100310), Other coffee (100330), Standard tea (100400), Rooibos tea (100410), Dairy smoothie (100230), Latte (100300), Cappuccino (100290), Type of milk used (100920), Added milk cereal (100890), Porridge (100770), Muesli (100800), Bran cereal (100840), Wholewheat cereal (100850), Oat crunch (100810), Plain cereal (100830), Sweetened cereal (100820), Other cereal (100860) | From 2.25 to < 2.5 | 3 |
| | | From 2 to < 2.25 | 4 |
| | | < 2 | 5 |
| **Discretionary foods** | Chocolate biscuit (102350), Chocolate covered biscuit (102340), Chocolate bar (102260), Chocolate sweets (102310), Chocolate raisins (102300), Dark chocolate (102290), Milk chocolate (102280), White chocolate (102270), Sweet biscuits (102360), Cakes (102190), Cheesecake (102220), Doughnut (102200), Fruitcake (102180), Danish pastry (102060), Sponge pudding (102210), Milk based pudding (102140), Other milk pudding (102150), Other desert (102230), Soya desert (102170), Sweets (102330), Diet sweets (102320), Other sweets (102380), Ice cream (102120), Fizzy drinks (100170), Squash (100180), Sugar added to tea (100490), Sugar added to coffee (100370), Sugar added to cereal (100900), Hot chocolate (100550), Pancake (102010), Scotch pancake (102020), Croissant (102050), Scone (102070), Crisp (102460), Cereal bar (102370) | ≥ 1.5 | 0 |
| | | From 1.375 to < 1.5 | 1 |
| | | From 1.25 to < 1.375 | 2 |
| | | From 1.12 to < 1.25 | 3 |
| | | From 1 to < 1.12 | 4 |
| | | < 1 | 5 |
| **Fluid** | Drinking water (100150), Instant coffee (100250), Filtered coffee (100270), Espresso (100310), Cappuccino (100290), Latte (100300), Other coffee type (100330), Decaffeinated coffee (100360), Standard tea (100400), Rooibos tea (100410), Green tea (100420), Herbal tea (100430), Other tea (100440), Decaffeinated tea (100470), Low calorie drink (100160), Squash (100180), Orange juice (100190), Grapefruit juice (100200), Pure fruit vegetable juice (100210), Fruit smoothie (100220), Dairy smoothie (100230), Type of milk used (100920), Milk (100520), Flavoured milk (100530) | From 0 to < 3 | 0 |
| | | From 3 to < 3.75 | 1 |
| | | From 3.75 to < 4.5 | 2 |
| | | From 4.5 to < 5.25 | 3 |
| | | From 5.25 to < 6 | 4 |
| | | ≥ 6 | 5 |

a; The final Eatwell Guide score for fish was created by averaging the Eatwell Guide score for oily fish and the EWG score for fish overall

**Supplementary Table S2 –** Comparison of baseline characteristics in the analytical cohort vs rest of cohort

| | **Analytical cohort (n = 156,764)** | **Rest of cohort (n = 345,455)** |
|---|---|---|
| **EWG adherence score** | 28.9 ± 8.4 | 28.1 ± 8.3 |
| **Age (years)** | 56.0 ± 8.0 | 56.8 ± 8.1 |
| **BMI (kg/m$^2$)** | 26.9 ± 4.5 | 27.7 ± 4.9 |
| **Sex (%)** | | |
| Females | 82788 (52.8) | 190423 (55.1) |
| Males | 73976 (47.2) | 155032 (44.9) |
| **Ethnicity (%)** | | |
| White | 150653 (96.1) | 321707 (93.1) |
| Non-white | 6111 (3.9) | 20264 (5.9) |
| **Education (%)** | | |
| Higher | 94133 (60.0) | 139286 (40.3) |
| Vocational | 18065 (11.5) | 44892 (13.0) |
| Upper secondary | 10015 (6.4) | 17200 (5.0) |
| Lower secondary | 22536 (14.4) | 60722 (17.6) |
| Other/prefer not to say | 12015 (7.7) | 78716 (22.8) |
| **Socioeconomic status (%)** | | |
| 1 (least deprived) | 34279 (21.9) | 67013 (19.4) |
| 2-4 | 97125 (62.0) | 203030 (58.8) |
| 5 (most deprived) | 25360 (16.1) | 74786 (21.6) |
| **Physical activity (%)** | | |
| Low | 28090 (17.9) | 43457 (12.6) |
| Moderate | 66202 (42.2) | 90272 (26.1) |
| High | 62472 (39.9) | 94576 (27.4) |
| **Smoking status (%)** | | |
| Never | 88859 (56.7) | 184519 (53.4) |
| Previous | 56106 (35.8) | 116844 (33.8) |
| Current | 11799 (7.5) | 41143 (11.9) |

Missing data for some variables from rest of cohort due to incomplete data available from UK Biobank dataset

**Supplementary Table S3 –** Linear regression analysis of associations between adherence to the Eatwell Guide and markers of adiposity

| Outcome | Beta | SE | P |
|---|---|---|---|
| BMI (n=156,764) | -0.032 | 0.001 | <0.001 |
| Waist circumference (n=156,942) | -0.088 | 0.003 | <0.001 |
| ABSI (n=156,764) | -0.005 | 0.0002 | <0.001 |
| Total body fat % (n=154,722) | -0.053 | 0.002 | <0.001 |
| Trunk body fat % (n=154,765) | -0.058 | 0.002 | <0.001 |

*ABSI is presented as sex-specific z score.*

**Supplementary table S4** - Logistic regression analysis of associations between adherence to the Eatwell Guide and odds of adiposity related outcomes

| Outcome | Low | | Moderate | | High | |
|---|---|---|---|---|---|---|
| | OR (95% CI) | P | OR (95% CI) | P | OR (95% CI) | P |
| BMI (n=156,764) | - | - | 0.91 (0.89-0.94) | <0.001 | 0.75 (0.73-0.77) | <0.001 |
| Waist circumference (n=156,942) | - | - | 0.91 (0.89-0.94) | <0.001 | 0.76 (0.74-0.78) | <0.001 |
| ABSI (n=156,764) | - | - | 0.90 (0.88-0.93) | <0.001 | 0.85 (0.82-0.87) | <0.001 |
| Total body fat % (n=154,722) | - | - | 0.92 (0.89-0.95) | <0.001 | 0.78 (0.75-0.80) | <0.001 |
| Trunk body fat % (n=154,765) | - | - | 0.90 (0.87-0.92) | <0.001 | 0.78 (0.75-0.80) | <0.001 |

*BMI split at 18.5 – 24.9 = healthy, 25+ = overweight/obesity. Waist circumference split at <94cm = healthy in men and <80cm = healthy in women. All other analyses split at group median.*

**Supplementary Table S5 –** Linear mixed model analysis of prospective associations between Eatwell Guide adherence tertile and score, and markers of weight by sex

| **Predictor (EWG tertile x time)** | **Sex** | | | | | | |
|---|---|---|---|---|---|---|---|
| | **Male** | | | **Female** | | | **Sex differences** |
| | Beta | Std. error | *P* value | Beta | Std. error | *P* value | *P* value |
| BMI | | | | | | | |
| Low (ref) | | | | | | | |
| Medium | -0.097 | 0.016 | <0.001 | -0.087 | 0.021 | <0.001 | 0.70 |
| High | -0.139 | 0.016 | <0.001 | -0.192 | 0.021 | <0.001 | 0.04 |
| Continuous | -0.007 | 0.001 | <0.001 | -0.011 | 0.001 | <0.001 | 0.02 |
| Waist circumference | | | | | | | |
| Low (ref) | | | | | | | |
| Medium | -0.264 | 0.061 | <0.001 | -0.287 | 0.073 | <0.001 | 0.83 |
| High | -0.353 | 0.062 | <0.001 | -0.483 | 0.071 | <0.001 | 0.18 |
| Continuous | -0.020 | 0.003 | <0.001 | -0.026 | 0.003 | <0.001 | 0.20 |
| ABSI | | | | | | | |
| Low (ref) | | | | | | | |
| Medium | -0.003 | 0.008 | 0.69 | 0.001 | 0.009 | 0.93 | 0.64 |
| High | 0.002 | 0.009 | 0.85 | -0.006 | 0.009 | 0.52 | 0.64 |
| Continuous | -0.0001 | 0.0004 | 0.90 | -0.0003 | 0.0004 | 0.47 | 0.75 |
| Total body fat % | | | | | | | |
| Low (ref) | | | | | | | |
| Medium | -0.130 | 0.032 | <0.001 | -0.076 | 0.041 | 0.07 | 0.31 |
| High | -0.237 | 0.033 | <0.001 | -0.266 | 0.040 | <0.001 | 0.57 |
| Continuous | -0.012 | 0.002 | <0.001 | -0.015 | 0.002 | <0.001 | 0.27 |
| Trunk body fat % | | | | | | | |
| Low (ref) | | | | | | | |
| Medium | -0.127 | 0.039 | <0.001 | -0.082 | 0.051 | 0.11 | 0.49 |
| High | -0.241 | 0.040 | <0.001 | -0.289 | 0.049 | <0.001 | 0.43 |
| Continuous | -0.127 | 0.002 | <0.001 | -0.017 | 0.002 | <0.001 | 0.19 |

Sex difference p-value corresponds to the statistical test of the three-way interaction term (EWG tertile × time × sex) in the combined model. N: BMI (male = 74,025, female = 82,898), Waist circumference (male = 74,123, female = 82,928), ABSI (male = 73,993, female = 82,841), Total body fat (male = 73,179, female = 82,057), Trunk fat percentage (male = 73,218, female = 82021).

**Supplementary Table S6** - Linear mixed model analysis of prospective associations between Eatwell Guide adherence tertile and score, and markers of weight by age

| Predictor (EWG tertile x time) | Age | | | | | | |
|---|---|---|---|---|---|---|---|
| | Younger | | | Older | | | Age differences |
| | Beta | Std. error | *P* value | Beta | Std. error | *P* value | *P* value |
| BMI | | | | | | | |
| Low (ref) | | | | | | | |
| Medium | -0.0816 | 0.0180 | <0.001 | -0.0488 | 0.0184 | <0.01 | 0.23 |
| High | -0.1534 | 0.0182 | <0.001 | -0.0911 | 0.0178 | <0.001 | 0.02 |
| Continuous | -0.0087 | 0.0009 | <0.001 | -0.0043 | 0.0008 | <0.001 | <0.001 |
| Waist circumference | | | | | | | |
| Low (ref) | | | | | | | |
| Medium | -0.2012 | 0.0620 | <0.01 | -0.1168 | 0.0725 | 0.11 | 0.40 |
| High | -0.3126 | 0.0626 | <0.001 | -0.1510 | 0.0704 | 0.03 | 0.10 |
| Continuous | -0.0191 | 0.0031 | <0.001 | -0.0063 | 0.0033 | 0.06 | <0.01 |
| ABSI | | | | | | | |
| Low (ref) | | | | | | | |
| Medium | 0.0074 | 0.0079 | 0.35 | 0.0082 | 0.0100 | 0.41 | 0.95 |
| High | 0.0102 | 0.0080 | 0.20 | 0.0126 | 0.0097 | 0.19 | 0.86 |
| Continuous | 0.0003 | 0.0004 | 0.39 | 0.0008 | 0.0005 | 0.07 | 0.43 |
| Total body fat % | | | | | | | |
| Low (ref) | | | | | | | |
| Medium | -0.1139 | 0.0347 | <0.001 | -0.0900 | 0.0389 | 0.02 | 0.66 |
| High | -0.2427 | 0.0350 | <0.001 | -0.2593 | 0.0379 | <0.001 | 0.74 |
| Continuous | -0.0136 | 0.0017 | <0.001 | -0.0132 | 0.0017 | <0.001 | 0.92 |
| Trunk body fat % | | | | | | | |
| Low (ref) | | | | | | | |
| Medium | -0.1295 | 0.0418 | <0.01 | -0.1047 | 0.0486 | 0.03 | 0.70 |
| High | -0.2730 | 0.0421 | <0.001 | -0.3055 | 0.0473 | <0.001 | 0.59 |
| Continuous | -0.0153 | 0.0020 | <0.001 | -0.0157 | 0.0022 | <0.001 | 0.90 |

Age split at the median (57) to categorise groups. Age difference p-value corresponds to the statistical test of the three-way interaction term (EWG tertile × time × age) in the combined model
N: BMI (younger = 81027, older = 75,871), waist circumference (younger = 81,097, older = 75,954), ABSI (younger = 80,993, older = 75,841), Total body fat (younger = 80,329, older = 74,907), Trunk fat (younger = 80,337, older = 74,902)

**Supplementary Table S7** - Linear mixed model analysis of prospective associations between Eatwell Guide adherence tertile and score, and markers of weight by engagement with physical activity

| Predictor (EWG tertile x time) | Physical activity | | | | | | | | | |
|---|---|---|---|---|---|---|---|---|---|---|
| | Low | | | Moderate | | | High | | | PA differences |
| | Beta | Std. error | *P* value | Beta | Std. error | *P* value | Beta | Std. error | *P* value | *P* value |
| BMI | | | | | | | | | | |
| Low (ref) | | | | | | | | | | |
| Medium | -0.0887 | 0.0333 | <0.01 | -0.0751 | 0.0196 | <0.001 | -0.0944 | 0.0203 | <0.001 | 0.87 |
| High | -0.1901 | 0.0346 | <0.001 | -0.1468 | 0.0196 | <0.001 | -0.1602 | 0.0196 | <0.001 | 0.43 |
| Continuous | -0.0099 | 0.0017 | <0.001 | -0.0087 | 0.0009 | <0.001 | -0.0077 | 0.0009 | <0.001 | 0.48 |
| Waist circumference | | | | | | | | | | |
| Low (ref) | | | | | | | | | | |
| Medium | -0.1923 | 0.1126 | 0.09 | -0.1978 | 0.0714 | <0.01 | -0.2424 | 0.0755 | <0.01 | 0.67 |
| High | -0.3072 | 0.1169 | <0.01 | -0.3336 | 0.0714 | <0.001 | -0.3369 | 0.0731 | <0.001 | 0.82 |
| Continuous | -0.0180 | 0.0057 | <0.01 | -0.0208 | 0.0034 | <0.001 | -0.0153 | 0.0035 | <0.001 | 0.58 |
| ABSI | | | | | | | | | | |
| Low (ref) | | | | | | | | | | |
| Medium | 0.0128 | 0.0140 | 0.35 | 0.0025 | 0.0094 | 0.78 | 0.0068 | 0.0102 | 0.50 | 0.73 |
| High | 0.0196 | 0.0145 | 0.18 | -0.0001 | 0.0094 | 0.93 | 0.0128 | 0.0098 | 0.19 | 0.75 |
| Continuous | 0.0005 | 0.0003 | 0.16 | 0.00001 | 0.0002 | 0.94 | 0.0003 | 0.0002 | 0.11 | 0.82 |
| Total body fat % | | | | | | | | | | |
| Low (ref) | | | | | | | | | | |
| Medium | -0.1715 | 0.0578 | <0.01 | -0.1032 | 0.0393 | <0.01 | -0.1450 | 0.0431 | <0.001 | 0.70 |
| High | -0.3427 | 0.0601 | <0.001 | -0.3148 | 0.0393 | <0.001 | -0.2827 | 0.0417 | <0.001 | 0.40 |
| Continuous | -0.0175 | 0.0029 | <0.001 | -0.0176 | 0.0019 | <0.001 | -0.0147 | 0.0020 | 0.001 | 0.41 |
| Trunk body fat % | | | | | | | | | | |
| Low (ref) | | | | | | | | | | |
| Medium | -0.1892 | 0.0698 | <0.01 | -0.1201 | 0.0481 | 0.01 | -0.1593 | 0.0527 | <0.01 | 0.72 |
| High | -0.3662 | 0.0726 | <0.001 | -0.3726 | 0.0481 | <0.001 | -0.3113 | 0.0511 | <0.001 | 0.52 |
| Continuous | -0.0190 | 0.0035 | <0.001 | -0.0207 | 0.0023 | <0.001 | -0.0162 | 0.0024 | <0.001 | 0.41 |

PA categorised as yes (engages in PA) and no (does not engage in PA). PA differences p-value corresponds to the statistical test of the three-way interaction term (EWG tertile × time × PA) in the combined model and represents high vs. low PA. N: BMI (Low = 28,133, Moderate = 66,265, High = 62,520), waist circumference (Low = 28.179, Moderate = 66,313, High = 62,559), ABSI (Low = 28,096, Moderate = 66,236, High = 62,502), Total body fat (Low =27,732, Moderate = 65,612, High = 61,892), Trunk fat (Low = 27,726, Moderate = 65,603, High =61,910)

**Supplementary Table S8** - Linear mixed model analysis of prospective associations between Eatwell Guide adherence tertile and score, and markers of weight by socio-economic status

| **Predictor (EWG tertile x time)** | **Socio-economic status** | | | | | | | | | |
|---|---|---|---|---|---|---|---|---|---|---|
| | **Least** | | | **Middle** | | | **Most** | | | **SES differences** |
| | Beta | Std. error | *P* value | Beta | Std. error | *P* value | Beta | Std. error | *P* value | *P* value |
| BMI | | | | | | | | | | |
| Low (ref) | | | | | | | | | | |
| Medium | -0.0610 | 0.0241 | 0.01 | -0.0623 | 0.0154 | <0.001 | -0.1370 | 0.037 | <0.001 | 0.10 |
| High | -0.1644 | 0.0241 | <0.001 | -0.1138 | 0.0153 | <0.001 | -0.2560 | 0.036 | <0.001 | 0.02 |
| Continuous | -0.0088 | 0.0012 | <0.001 | -0.0065 | 0.0008 | <0.001 | -0.0134 | 0.0008 | <0.001 | 0.03 |
| Waist circumference | | | | | | | | | | |
| Low (ref) | | | | | | | | | | |
| Medium | -0.128 | 0.0902 | 0.16 | -0.176 | 0.0600 | <0.01 | -0.407 | 0.138 | <0.01 | 0.07 |
| High | -0.333 | 0.0899 | <0.001 | -0.201 | 0.0594 | <0.001 | -0.650 | 0.136 | <0.001 | 0.046 |
| Continuous | -0.0169 | 0.0043 | <0.001 | -0.0122 | 0.0029 | <0.001 | -0.0325 | 0.0064 | <0.001 | 0.04 |
| ABSI | | | | | | | | | | |
| Low (ref) | | | | | | | | | | |
| Medium | -0.0002 | 0.0123 | 0.99 | 0.0132 | 0.0079 | 0.09 | -0.0064 | 0.0171 | 0.71 | 0.78 |
| High | 0.0022 | 0.0122 | 0.86 | 0.0176 | 0.0078 | 0.02 | -0.0042 | 0.0169 | 0.80 | 0.79 |
| Continuous | 0.0004 | 0.0006 | 0.55 | 0.0007 | 0.0004 | 0.049 | -0.0002 | 0.0008 | 0.85 | 0.65 |
| Total body fat % | | | | | | | | | | |
| Low (ref) | | | | | | | | | | |
| Medium | -0.1075 | 0.0506 | 0.03 | -0.1246 | 0.0328 | <0.001 | -0.1186 | 0.0768 | 0.12 | 0.86 |
| High | -0.2022 | 0.0503 | <0.001 | -0.2791 | 0.0325 | <0.001 | -0.4477 | 0.0759 | <0.001 | 0.01 |
| Continuous | -0.0106 | 0.0024 | <0.001 | -0.0155 | 0.0016 | <0.001 | -0.0213 | 0.0036 | <0.001 | 0.01 |
| Trunk body fat % | | | | | | | | | | |
| Low (ref) | | | | | | | | | | |
| Medium | -0.1075 | 0.0506 | 0.03 | -0.1246 | 0.0328 | <0.001 | -0.1186 | 0.7677 | 0.12 | 0.91 |
| High | -0.2022 | 0.0503 | <0.001 | -0.2791 | 0.0325 | <0.001 | -0.4477 | 0.0759 | <0.001 | 0.01 |
| Continuous | -0.0117 | 0.0030 | <0.001 | -0.0181 | 0.0019 | <0.001 | -0.0229 | 0.0043 | <0.001 | 0.03 |

SES categorised based on Townsend deprivation index split into tertiles. SES differences p-value corresponds to the statistical test of the three-way interaction term (EWG tertile × time × SES) in the combined model and represents least vs. most SES. N: BMI (least = 41,010, middle = 117,555, most = 31,278), waist circumference (least = 34,329, middle = 97,298, most = 25,424), ABSI (least = 34,296, middle = 97,168, most = 25,370), Total body fat (least = 33,994, middle = 96,173, most = 25,069), Trunk fat (least = 34,000 , middle = 96,182, most = 25,057)

**Supplementary Table S9** - Linear mixed model analysis of prospective associations between Eatwell Guide adherence tertile and score, and markers of weight by polygenic risk score for obesity

| **Predictor (EWG tertile x time)** | **PRS** | | | | | | |
|---|---|---|---|---|---|---|---|
| | **Low** | | | **High** | | | **PRS differences** |
| | Beta | Std. error | *P* value | Beta | Std. error | *P* value | *P* value |
| BMI | | | | | | | |
| Low (ref) | | | | | | | |
| Medium | -0.0685 | 0.0153 | <0.001 | -0.0811 | 0.0234 | <0.001 | 0.63 |
| High | -0.1339 | 0.0152 | <0.001 | -0.1642 | 0.0232 | <0.001 | 0.25 |
| Continuous | -0.0077 | 0.0007 | <0.001 | -0.0083 | 0.0011 | <0.001 | 0.62 |
| Waist circumference | | | | | | | |
| Low (ref) | | | | | | | |
| Medium | -0.1735 | 0.0589 | <0.01 | -0.0811 | 0.0234 | <0.001 | 0.90 |
| High | -0.2655 | 0.0584 | <0.001 | -0.1642 | 0.0232 | <0.001 | 0.53 |
| Continuous | -0.0156 | 0.0028 | <0.001 | -0.1701 | 0.0038 | <0.001 | 0.79 |
| ABSI | | | | | | | |
| Low (ref) | | | | | | | |
| Medium | 0.0063 | 0.0081 | 0.44 | 0.0094 | 0.0010 | 0.34 | 0.78 |
| High | 0.0110 | 0.0080 | 0.17 | 0.0106 | 0.0010 | 0.28 | 0.99 |
| Continuous | 0.0005 | 0.0003 | 0.16 | 0.0005 | 0.0005 | 0.28 | 0.96 |
| Total body fat % | | | | | | | |
| Low (ref) | | | | | | | |
| Medium | -0.1151 | 0.0323 | <0.001 | 0.0098 | 0.0434 | 0.02 | 0.76 |
| High | -0.2827 | 0.0322 | <0.001 | 0.2747 | 0.0429 | <0.001 | 0.89 |
| Continuous | -0.0160 | 0.0016 | <0.001 | -0.0134 | 0.0021 | <0.001 | 0.44 |
| Trunk body fat % | | | | | | | |
| Low (ref) | | | | | | | |
| Medium | -0.1236 | 0.0399 | <0.01 | -0.1098 | 0.0528 | 0.03 | 0.80 |
| High | -0.3229 | 0.0396 | <0.001 | -0.3061 | 0.0522 | <0.001 | 0.81 |
| Continuous | -0.0181 | 0.0019 | <0.001 | -0.0161 | 0.0025 | <0.001 | 0.52 |

PRS split at <0 and >0 to categorise groups. PRS difference p-value corresponds to the statistical test of the three-way interaction term (EWG tertile × time × age) in the combined model N: BMI (low = 90,733, high = 62,872), waist circumference (low = 90,820, high = 62,932), ABSI (low = 90,703, high = 62,843), Total body fat (low = 89,814, high = 62,178), Trunk fat (low = 89,823, high = 62,171)

**Supplementary Table S10 -** Sensitivity analysis for the linear mixed model analysis of prospective associations between Eatwell Guide adherence tertile and score, and markers of weight by polygenic risk score for obesity, but with PRS split into tertiles

| Predictor (EWG tertile x time) | PRS | | | | | | | | | |
|---|---|---|---|---|---|---|---|---|---|---|
| | **Low** | | | **Moderate** | | | **High** | | | **PRS differences** |
| | Beta | Std. error | *P* value | Beta | Std. error | *P* value | Beta | Std. error | *P* value | *P* value |
| BMI | | | | | | | | | | |
| Low (ref) | | | | | | | | | | |
| Medium | -0.0824 | 0.0243 | <0.001 | -0.0651 | 0.0165 | <0.001 | -0.0894 | 0.0363 | 0.01 | 0.83 |
| High | -0.1468 | 0.0241 | <0.001 | -0.1393 | 0.0163 | <0.001 | -0.1657 | 0.0361 | <0.001 | 0.64 |
| Continuous | -0.0083 | 0.0012 | <0.001 | -0.0078 | -0.0007 | <0.001 | -0.0083 | 0.0018 | <0.001 | 0.99 |
| Waist circumference | | | | | | | | | | |
| Low (ref) | | | | | | | | | | |
| Medium | -0.2146 | 0.0985 | 0.03 | -0.1386 | 0.0604 | 0.02 | -0.2670 | 0.1176 | 0.02 | 0.71 |
| High | -0.2625 | 0.0978 | <0.01 | -0.2970 | 0.0600 | <0.001 | -0.3013 | 0.1170 | 0.01 | 0.79 |
| Continuous | -0.0149 | 0.0047 | <0.01 | -0.0164 | 0.0029 | <0.001 | -0.0173 | 0.0057 | <0.01 | 0.74 |
| ABSI | | | | | | | | | | |
| Low (ref) | | | | | | | | | | |
| Medium | 0.0117 | 0.0139 | 0.40 | 0.0072 | 0.0080 | 0.37 | 0.0038 | 0.0142 | 0.79 | 0.74 |
| High | 0.0212 | 0.0138 | 0.12 | 0.0052 | 0.0080 | 0.52 | 0.0162 | 0.0141 | 0.25 | 0.82 |
| Continuous | 0.0011 | 0.0007 | 0.10 | 0.0003 | 0.0004 | 0.38 | 0.0005 | 0.0007 | 0.51 | 0.52 |
| Total body fat % | | | | | | | | | | |
| Low (ref) | | | | | | | | | | |
| Medium | -0.1282 | 0.0542 | 0.02 | -0.1121 | 0.033 | <0.001 | -0.0760 | 0.0635 | 0.23 | 0.55 |
| High | -0.2790 | 0.0542 | <0.001 | -0.2888 | 0.033 | <0.001 | -0.2528 | 0.0630 | <0.001 | 0.75 |
| Continuous | -0.0171 | 0.0026 | <0.001 | -0.0155 | 0.0016 | <0.001 | -0.0123 | 0.0031 | <0.001 | 0.23 |
| Trunk body fat % | | | | | | | | | | |
| Low (ref) | | | | | | | | | | |
| Medium | -0.1257 | 0.0679 | 0.06 | -0.1264 | 0.0407 | <0.01 | -0.0942 | 0.0770 | 0.22 | 0.77 |
| High | -0.3057 | 0.0673 | <0.001 | -0.3362 | 0.0404 | <0.001 | -0.2665 | 0.0762 | <0.001 | 0.69 |
| Continuous | -0.1915 | 0.0032 | <0.001 | -0.1800 | 0.0019 | <0.001 | -0.0131 | 0.0037 | <0.001 | 0.22 |

PRS split into quintiles and then collapsed into tertiles (low = 1; moderate = 2,3,4; high = 5) to categorise groups. PRS difference p-value corresponds to the statistical test of the three-way interaction term (EWG tertile × time × age) in the combined model N: BMI (low = 30,876, moderate = 92,155, high = 30,574), waist circumference (low = 30,905, moderate = 92,250, high = 30,597), ABSI (low = 30,864, moderate = 92,120, high = 30,562), Total body fat (low = 30,571, moderate = 91,210, high = 30,211), Trunk fat (low = 30,576, moderate = 91,216, high = 30,202

**Supplementary Table S11 -** Sensitivity analysis of prospective association with Eatwell Guide adherence score and markers of weight, but with participants with a minimum of two dietary reports

| Predictor (EWG score x time) | Beta | Std.Error | P-value |
|---|---|---|---|
| BMI (n = 102,013) | -0.0076 | 0.0007 | <0.001 |
| Waist circumference (n = 102,088) | -0.0144 | 0.0026 | <0.001 |
| ABSI (n = 101,967) | 0.0006 | 0.0003 | 0.08 |
| Total body fat % (n = 100,998) | -0.0162 | 0.0014 | <0.001 |
| Trunk body fat % (n = 101,005) | -0.0186 | 0.0018 | <0.001 |

**Supplementary Table S12 -** Sensitivity analysis of prospective association with Eatwell Guide adherence score and markers of weight, but excluding participants with extreme energy intakes

| Predictor (EWG score x time) | Beta | Std.Error | P-value |
|---|---|---|---|
| BMI (n = 155,601) | -0.0082 | 0.0006 | <0.001 |
| Waist circumference (n = 155,753) | -0.0161 | 0.0023 | <0.001 |
| ABSI (n = 155,537) | 0.0006 | 0.0003 | 0.04 |
| Total body fat % (153,960) | -0.0155 | 0.0012 | <0.001 |
| Trunk body fat % (153,960) | -0.0176 | 0.0015 | <0.001 |

**Supplementary Table S13 -** Sensitivity analysis of prospective association with Eatwell Guide adherence score and markers of weight, but removing energy intake as a covariate from the model

| Predictor (EWG score x time) | Beta | Std.Error | P-value |
|---|---|---|---|
| BMI (n = 156,898) | -0.0080 | 0.0006 | <0.001 |
| Waist circumference (n = 157,051) | -0.0162 | 0.0023 | <0.001 |
| ABSI (n = 156,834) | 0.0005 | 0.0002 | 0.06 |
| Total body fat % (155,236) | -0.0151 | 0.0012 | <0.001 |
| Trunk body fat % (153,960) | -0.0171 | 0.0015 | <0.001 |

**Supplementary Table S14** - Sensitivity analysis of prospective association with Eatwell Guide adherence score and BMI, with one component sequentially removed from the score

| **Predictor (EWG score x time)** | **Beta** | **Std.Error** | **P-value** |
|---|---|---|---|
| EWG score: | | | |
| Minus starchy carbohydrate | -0.0085 | 0.0007 | <0.001 |
| Minus wholegrains | -0.0076 | 0.0007 | <0.001 |
| Minus red meat | -0.0082 | 0.0007 | <0.001 |
| Minus fish | -0.0081 | 0.0007 | <0.001 |
| Minus white meat | -0.0100 | 0.0007 | <0.001 |
| Minus fruit and veg | -0.0071 | 0.0007 | <0.001 |
| Minus dairy | -0.0080 | 0.0006 | <0.001 |
| Minus beans and pulses | -0.0082 | 0.0007 | <0.001 |
| Minus nuts | -0.0083 | 0.0007 | <0.001 |
| Minus eggs | -0.0087 | 0.0007 | <0.001 |
| Minus discretionary food | -0.0073 | 0.0007 | <0.001 |
| Minus fluid | -0.0077 | 0.0007 | <0.001 |

**Supplementary table S15** - Sensitivity analysis of prospective association with Eatwell Guide adherence score and waist circumference, with one component sequentially removed from the score

| **Predictor (EWG score x time)** | **Beta** | **Std.Error** | **P-value** |
|---|---|---|---|
| EWG score | | | |
| Minus starchy carbohydrate | -0.0174 | 0.0024 | <0.001 |
| Minus wholegrains | -0.0145 | 0.0024 | <0.001 |
| Minus red meat | -0.0175 | 0.0024 | <0.001 |
| Minus fish | -0.0167 | 0.0024 | <0.001 |
| Minus white meat | -0.0187 | 0.0024 | <0.001 |
| Minus fruit and veg | -0.0154 | 0.0024 | <0.001 |
| Minus dairy | -0.0164 | 0.0024 | <0.001 |
| Minus beans and pulses | -0.0157 | 0.0024 | <0.001 |
| Minus nuts | -0.0163 | 0.0024 | <0.001 |
| Minus eggs | -0.0171 | 0.0024 | <0.001 |
| Minus discretionary food | -0.0147 | 0.0024 | <0.001 |
| Minus fluid | -0.0161 | 0.0024 | <0.001 |

**Supplementary Table S16** - Sensitivity analysis of prospective association with Eatwell Guide adherence score and ABSI, with one component sequentially removed from the score

| **Predictor (EWG score x time)** | **Beta** | **Std.Error** | **P-value** |
|---|---|---|---|
| EWG score | | | |
| Minus starchy carbohydrate | 0.0005 | 0.0003 | 0.11 |
| Minus wholegrains | 0.0006 | 0.0003 | 0.06 |
| Minus red meat | 0.0004 | 0.0003 | 0.17 |
| Minus fish | 0.0004 | 0.0003 | 0.13 |
| Minus white meat | 0.0007 | 0.0003 | 0.02 |
| Minus fruit and veg | 0.0003 | 0.0003 | 0.30 |
| Minus dairy | 0.0006 | 0.0003 | 0.048 |
| Minus beans and pulses | 0.0007 | 0.0003 | 0.02 |
| Minus nuts | 0.0006 | 0.0003 | 0.047 |
| Minus eggs | 0.0007 | 0.0003 | 0.02 |
| Minus discretionary food | 0.0006 | 0.0003 | 0.04 |
| Minus fluid | 0.0005 | 0.0003 | 0.14 |

**Supplementary Table S17 -** Sensitivity analysis of prospective association with Eatwell Guide adherence score and body fat percentage, with one component sequentially removed from the score

| **Predictor (EWG score x time)** | **Beta** | **Std.Error** | **P-value** |
|---|---|---|---|
| EWG score | | | |
| Minus starchy carbohydrate | -0.0154 | 0.0013 | <0.001 |
| Minus wholegrains | -0.0156 | 0.0013 | <0.001 |
| Minus red meat | -0.0153 | 0.0013 | <0.001 |
| Minus fish | -0.0160 | 0.0013 | <0.001 |
| Minus white meat | -0.0174 | 0.0013 | <0.001 |
| Minus fruit and veg | -0.0134 | 0.0013 | <0.001 |
| Minus dairy | -0.0156 | 0.0012 | <0.001 |
| Minus beans and pulses | -0.0161 | 0.0013 | <0.001 |
| Minus nuts | -0.0160 | 0.0013 | <0.001 |
| Minus eggs | -0.0160 | 0.0013 | <0.001 |
| Minus discretionary food | -0.0138 | 0.0013 | <0.001 |
| Minus fluid | -0.0139 | 0.0013 | <0.001 |

**Supplementary Table S18** - Sensitivity analysis of prospective association with Eatwell Guide adherence score and trunk fat percentage, with one component sequentially removed from the score

| **Predictor (EWG score x time)** | **Beta** | **Std.Error** | **P-value** |
|---|---|---|---|
| EWG score | | | |
| Minus starchy carbohydrate | -0.0175 | 0.0016 | <0.001 |
| Minus wholegrains | -0.0182 | 0.0016 | <0.001 |
| Minus red meat | -0.0172 | 0.0016 | <0.001 |
| Minus fish | -0.0184 | 0.0016 | <0.001 |
| Minus white meat | -0.0200 | 0.0016 | <0.001 |
| Minus fruit and veg | -0.0150 | 0.0016 | <0.001 |
| Minus dairy | -0.0179 | 0.0015 | <0.001 |
| Minus beans and pulses | -0.0184 | 0.0016 | <0.001 |
| Minus nuts | -0.0184 | 0.0016 | <0.001 |
| Minus eggs | -0.0182 | 0.0016 | <0.001 |
| Minus discretionary food | -0.0155 | 0.0016 | <0.001 |
| Minus fluid | -0.0154 | 0.0016 | <0.001 |
